# Fully automatic segmentation of brain lacunas resulting from resective surgery using a 3D deep learning model

**DOI:** 10.1101/2023.11.16.23298572

**Authors:** Raphael Fernandes Casseb, Brunno Machado de Campos, Wallace Souza Loos, Marcelo Eduardo Ramos Barbosa, Marina Koutsodontis Machado Alvim, Gabriel Chagas Lutfala Paulino, Francesco Pucci, Samuel Worrell, Roberto Medeiros de Souza, Lara Jehi, Fernando Cendes

## Abstract

The rapid and constant development of deep learning (DL) strategies is pushing forward the quality of object segmentation in images from diverse fields of interest. In particular, these algorithms can be very helpful in delineating brain abnormalities (lesions, tumors, lacunas, etc), enabling the extraction of information such as volume and location, that can inform doctors or feed predictive models. Here, we describe ResectVol DL, a fully automatic tool developed to segment resective lacunas in brain images of patients with epilepsy. ResectVol DL relies on the nnU-Net framework that leverages the 3D U-Net deep learning architecture. T1-weighted MRI datasets from 120 patients (57 women; 31.5 ± 15.9 years old at surgery) were used to train (n=78) and test (n=48) our tool. Manual segmentations were carried out by five different raters and were considered as ground truth for performance assessment. We compared ResectVol DL with two other fully automatic methods: ResectVol 1.1.2 and DeepResection, using the Dice similarity coefficient (DSC), Pearson’s correlation coefficient, and relative difference to manual segmentation. ResectVol DL presented the highest median DSC (0.92 vs. 0.78 and 0.90), the highest correlation coefficient (0.99 vs. 0.63 and 0.94), and the lowest median relative difference (9 vs. 44 and 12 %). Overall, we demonstrate that ResectVol DL accurately segments brain lacunas, which has the potential to assist in the development of predictive models for postoperative cognitive and seizure outcomes.

## 1. Introduction

Epilepsy is a neurological disease with profound impacts on quality of life, morbidity, and mortality. About 30-40 % of patients do not respond to treatment with medication (1,2) and can be referred to surgical intervention, especially if structural alterations are detected (3,4). Many factors impact surgical outcome and long-term seizure freedom is still limited, ranging from 40 to 80 % (5,6)

Studies have tried to determine the factors leading to surgical success (7–9) and nomograms were created showing promising results for individualizing outcome prediction (5,10). However, most surgical outcome studies still capture the surgical procedure under broad categorical classifications with resolution limited to the lobe of resection (temporal/external temporal, hippocampal sparing versus resecting).

Characterization of the lacuna can be informative to those predictive models by providing volume and location of extracted brain tissue. Due to the laborious and time-consuming nature of the manual annotation required to characterize resections, initiatives to automate this task increased in the last years (11–13). Hence, the purpose of this study is twofold: (i) to introduce ResectVol DL, a fully automatic method to segment brain lacunas, based on the 3D U-Net architecture (14); and (ii) to compare ResectVol DL against two other methods: ResectVol (version 1.1.2), a rule-based processing pipeline (12), and DeepResection, which is based on a 2D U-Net architecture (13).

## 2. Material and Methods

This study was conducted with approval from the Cleveland Clinical Foundation Institutional Review Board. Informed consent was waived due to the retrospective nature of the data collection.

### 2.1. Subjects and image datasets

Image datasets from 125 patients were retrospectively selected from the Cleveland Clinic Epilepsy Center and the University of Campinas. We included subjects with temporal (n=45) and extratemporal lobe epilepsy (n=75), and volunteers with no brain lesions or resective intervention (n=5) to serve as controls for false discoveries (no-surgery data). Only resective surgeries were included.

### 2.2. Manual segmentation

Five raters trained in neuroanatomy performed the manual segmentation of the surgical lacunas using MRIcron (15). Forty-eight image datasets were segmented by either two (n=42) or three (n=6) raters and were analyzed for inter-rater agreement.

### 2.3. Automatic Segmentation

To perform the automatic segmentation, we created ResectVol DL, which is based on the nnU-Net deep learning framework (16). nnU-Net derives from the U-Net (14), a popular deep learning architecture composed of an encoder, a decoder, and skip connections that concatenate features from the encoder to the decoder part of the network. Each processing stage of the encoding and decoding streams has two blocks of operation comprising steps of convolution, intensity normalization, and linear regularization (**Fig. 1**). Data augmentation strategies included rotation, scaling, addition of Gaussian noise, Gaussian blurring, changes to brightness and contrast, simulation of low resolution, and mirroring. The model training was carried out with five cross-validation folds and 1000 epochs for each fold.

**Figure 1.**
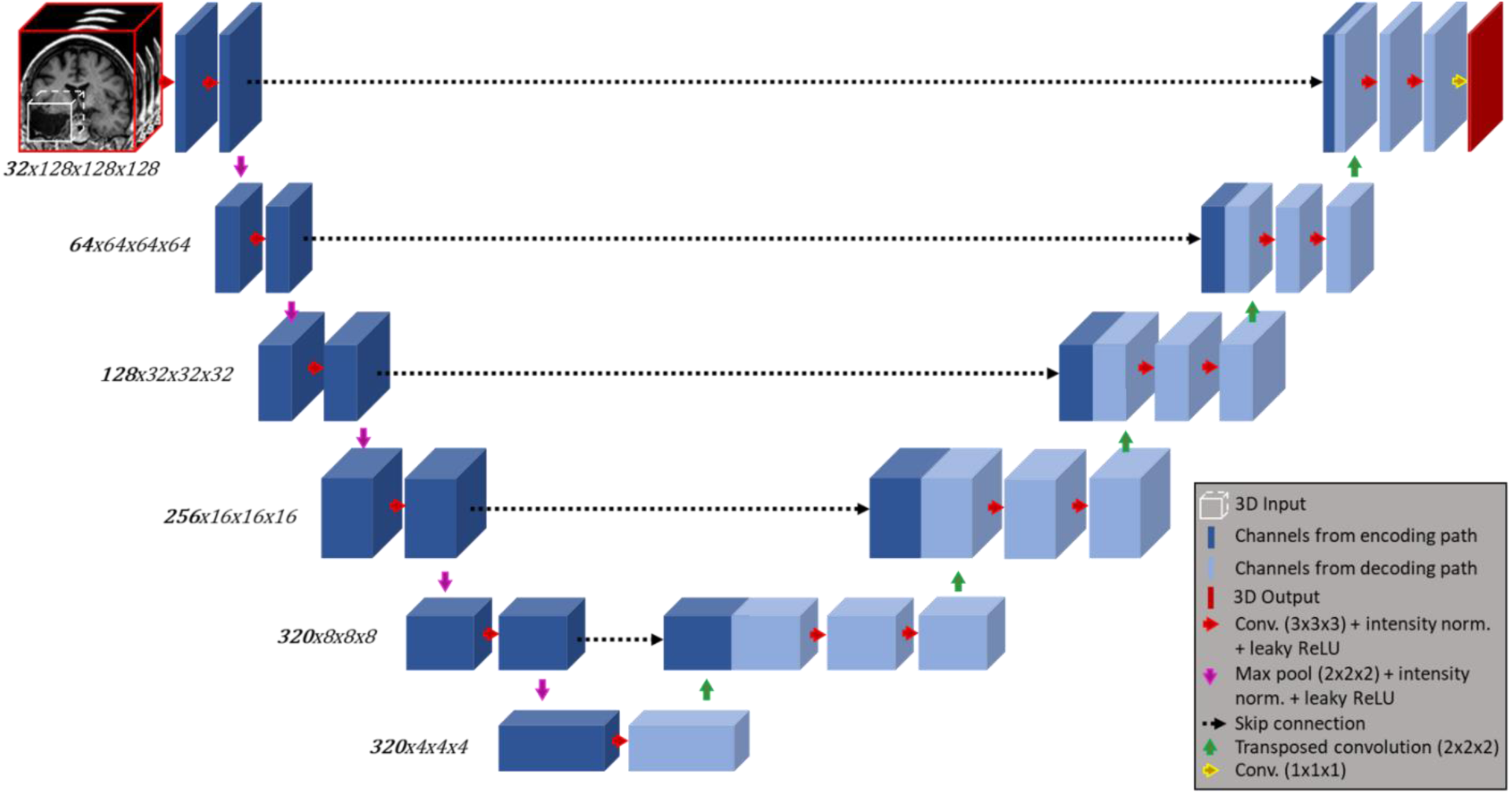
Representation of the 3D nnU-Net architecture. The number of channels and resolution at each of the six stages are shown on the left (eg: there are 320 channels and a resolution of 4x4x4 in the feature map of the sixth stage). Contiguous blocks after the green arrows represent the concatenation of channels from the lower stage of the decoding branch (light blue) with channels at the same stage from the encoding branch (dark blue).

To compare our algorithm against other freely available methods, we also performed the lacuna prediction with two other tools: ResectVol version 1.1.2 (available at lniunicamp.com/resectvol), a rule-based algorithm previously developed by our group that does not employ deep learning methods (12); and the DeepResection tool (13) (available at github.com/penn-cnt/DeepResection) which uses a set of three 2D U-Nets, each in one anatomical plane, followed by voxel-wise majority voting. In ResectVol 1.1.2, users must provide the pre- and postoperative images to run the analysis, whereas in DeepResection users are required to provide the postoperative image only.

### 2.4. Computational Resources

All automatic processing was performed on a 3.6 GHz Intel(R) Core(TM) i7-12700KF CPU with 32-GB RAM, a 48-GB NVIDIA RTX A6000, and a 20.04.1 Ubuntu operational system.

### 2.5. Performance Assessment

The Dice similarity coefficient (DSC) (17) was used as an indirect measure of overlap between manual and automatic segmentations. We also calculated the volume obtained from the manual and the automatic segmentations, and estimated Pearson’s correlation and the volume difference percentage (manual volume as reference) between them.

Additionally, to investigate if there were differences associated with lacuna size, we split lacunas into two groups (large and small) based on their median volume and compared the DSCs. Regarding the inter-rater segmentation performance, we calculated the DSC for the 48 images that were manually segmented more than once.

## 3. Results

### 3.1. Subjects

One hundred and twenty patient volunteers (57 women; 31.5 ± 15.9 years old at surgery) had temporal (n=45; 23 women; 37.7 ± 13.8 years old) and extratemporal (n=75; 34 women; 27.8 ± 16.0 years old) epilepsy, whereas five subjects (2 women; 29.0 ± 4.5 years old) were included as controls who were not submitted to any brain surgery. Nearly all postoperative images were acquired 5 months after surgery (median: 6.2; Q1-Q3: 6.0 - 8.0; range: .2 - 63.9), to avoid necrotic tissue that could be misleading in the segmentation process. Seventy-three patients (∼61 %) were seizure-free (Engel Ia) after surgery.

### 3.2. Imaging protocol

T1-weighted anatomical MR images were obtained in a total of eight different MRI systems (1.5 and 3 T) from two different vendors (Siemens and Philips). Acquisition parameters and image quality varied, benefiting the robustness of the performance assessment. Parameters can be summarized in the following ranges: voxel size = 0.41 × 0.45 × 0.41 to 1 × 2 × 1 mm³; TR = 7 to 2200 ms; TE = 2.3 to 46 ms; and image matrix = 180 x 96 to 512 x 512.

### 3.3. Manual segmentation

All the images containing brain lacunas (n=120) were manually segmented at least once. Three of the five raters timed the duration of manual delineation in a subset of images yielding the approximated median times of: 114 min (MERB, n=31); 70 min (RFC, n=14); 54 min (GCLP, n=19). Forty-eight images were segmented by two different raters at least, and the median DSC was .88 (Q1-Q3: .78 - .91; range: .00 - .97). We visually selected the best manual mask in these cases to compose the final set of 120 manual masks used as ground truth in the subsequent analyses.

### 3.4. ResectVol DL

Seventy-two images were used to train the nnU-Net architecture. We found a median DSC of .92 (Q1- Q3: .88 - .94) and .91 (Q1-Q3: .86 - .94) for the test and the cross-validation set, respectively (**Table 1**).

**Table 1.** Dice Similarity Coefficient (DSC), correlation, and relative difference obtained with each method for the test set.

|  |  | *ResectVol DL* | *ResectVol 1.1.2* | *DeepResection* |
| --- | --- | --- | --- | --- |
| **All data** | median DSC (Q1-Q3) | **.92 (.88 - .94)** | .78 (.65 - .84) | .07 (0 - .89)* |
|  | range | 0 - .96 | 0 - .90 | 0 - .95* |
|  | correlation (p-val) | .99 (< .001) | .63 (< .001) | ---- |
|  | median rel. diff. (Q1-Q3) | 9 (4 - 15) % | 44 (29 - 75) % | ---- |
| **Temporal** | median DSC (Q1-Q3) | **.93 (.89 - .95)** | .79 (.75 - .86) | .90 (.86 - .92) |
|  | range | .85 - .96 | .21 - .90 | .20 - .95 |
|  | correlation (p-val) | .97 (< .001) | .72 (< .001) | .94 (< .001) |
|  | median rel. diff. (Q1-Q3) | 8 (4 - 11) % | 45 (30 - 65) % | 12 (6 - 20) % |
| **Extra-temporal** | median DSC (Q1-Q3) | **.92 (.88 - .94)** | .73 (.61 - .79) | 0 (0 - 0)* |
|  | range | 0 - .96 | 0 - .89 | 0 - .08* |
|  | correlation (p-val) | .99 (< .001) | .62 (.001) | ---- |
|  | median rel. diff. (Q1-Q3) | 10 (4 - 20) % | 37 (15 - 84) % | ---- |
| **No-surgery data** | **5 cases** | **0** | **5** | **0** |

\* *DeepResection* authors do not recommend applying it to extra-temporal cases. These values are just reported for completeness. The associated correlation values, however, are not displayed due to the impossibility related to data distribution (see **Fig. 2A**, bottom plot).

For 47 of the 48 test images, ResectVol DL was able to correctly identify the lacuna (DSC ≥ .612) and it failed for only one extratemporal case – the smallest lacuna in the sample. The correlation between the manual and the automatic volumes was r(46) = .99 (p < .001), with a median relative difference of 9 % (Q1-Q3: 4 – 15 %) (**Table 1** and **Fig. 2**). ResectVol DL correctly identified the absence of lacunas in the five control datasets. The prediction for each image took, on average, 41.7 seconds.

**Figure 2.**
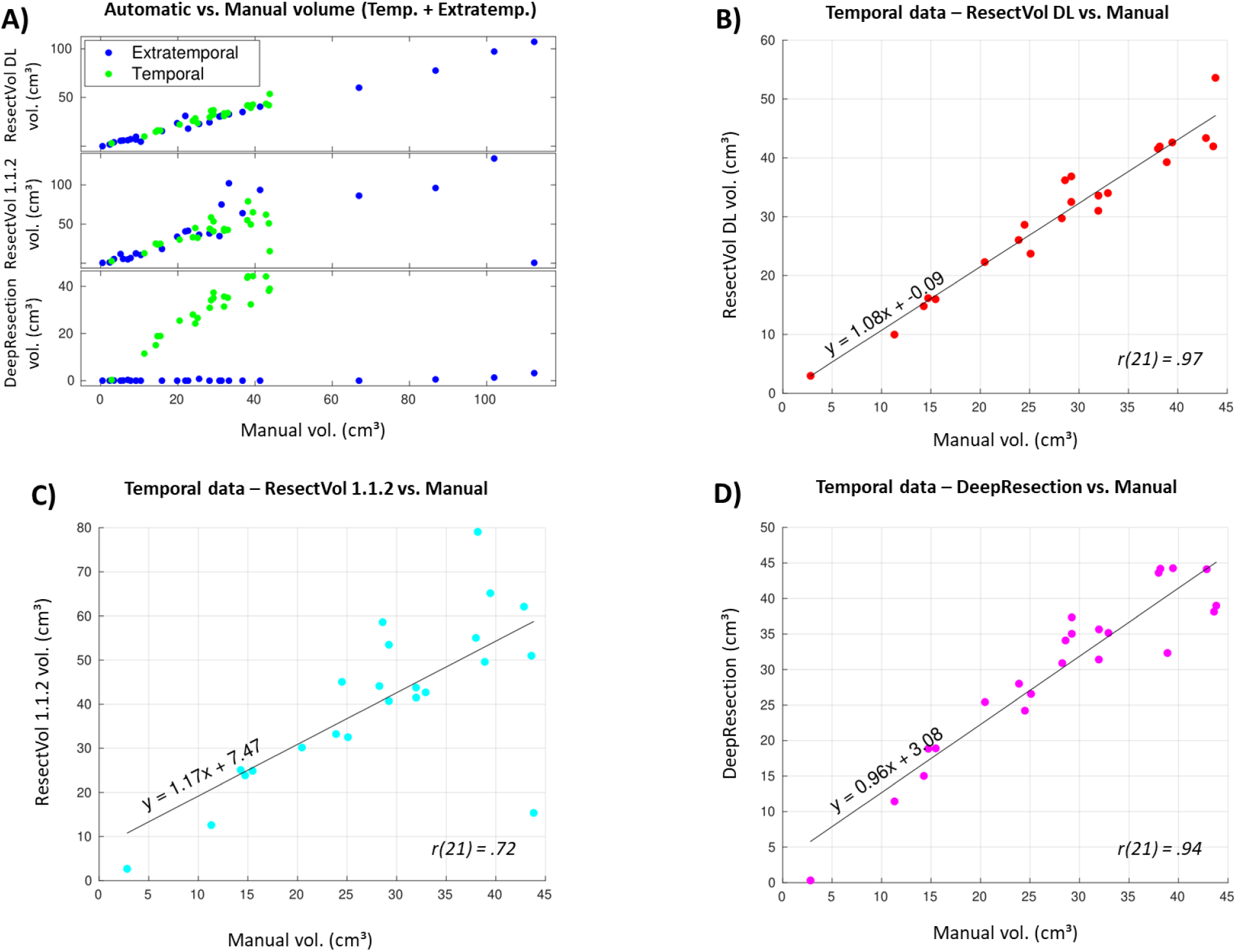
Scatter plots for (**A**) all data (temporal + extratemporal cases) and for (**B**-**D**) temporal data only between each segmentation method and manual delineation. Plots **B**-**D** include the linear fit, the equation that best adjusts to the data, and the correlation value r.

### 3.5. Performance of other approaches

The test set of 53 images (48 lacunas and five no-surgery controls) was also processed by ResectVol 1.1.2 and DeepResection.

ResectVol 1.1.2 requires no pre-processing of the images, although the origin of image space (x=0, y=0, z=0) must not be too far off the anterior commissure, in which case reorienting is necessary. It took on average nine minutes and 17 seconds to process each dataset composed of the pre- and the postoperative images. We found a median DSC = .78 (Q1-Q3: .65 - .84), a correlation between the manual and automatic volumes of r(46) = .63 (p < .001), and a median relative difference of 44 % (Q1-Q3: 29 – 75 %). For the five control images, it incorrectly found a lacuna in all five cases (volume range: .3 - 4.2 cm³) (**Table 1** and **Fig. 2**).

Like the other tools, DeepResection does not require any image preprocessing. The authors do recommend, however, that images are in Left-Anterior-Superior orientation because predictions can be inaccurate otherwise. We used *fslorient* function from the FMRIB Software Library (FSL) (18) to perform the reorientation. The available version of DeepResection is only intended for temporal resections, thus results for this tool are mostly restricted to this brain region. Temporal lacuna predictions overlapped with the manual segmentation in all cases, yielding a DSC = .90 (.86 - .92). Median correlation and relative difference were r(46) = .94 (p < .001) and 12 % (Q1-Q3: 6 – 20 %), respectively (**Table 1** and **Fig. 2**).

**Fig. 3** exhibits the best, median, and worst DSC-associated cases for all tools.

**Fig. 3.**
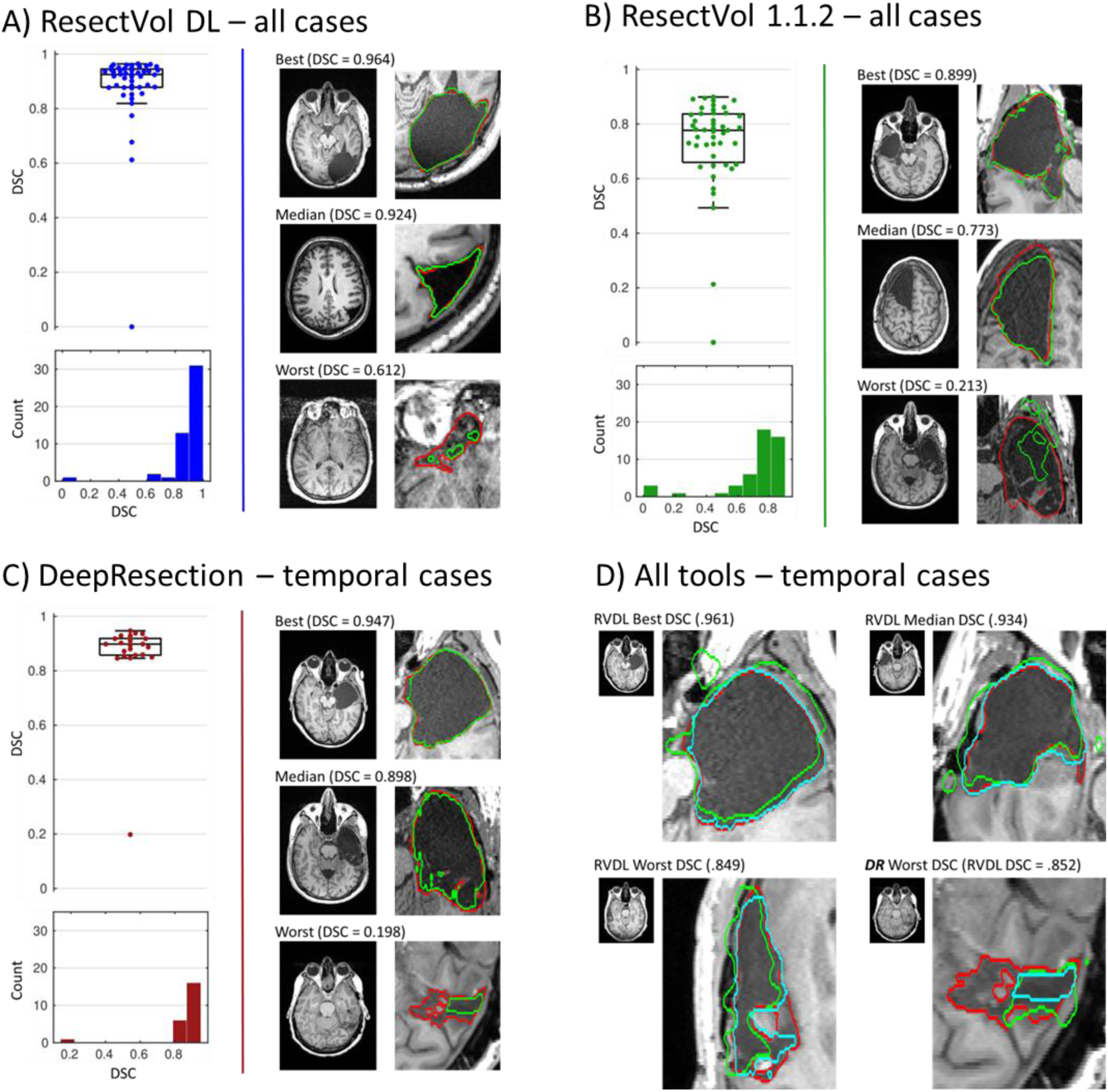
Boxplots, histograms, and noteworthy cases for (**A**) ResectVol DL (RV2), (**B**) ResectVol 1.1.2 (RV112), and (**C**) DeepResection (DR), along with Dice similarity coefficient (DSC). Red and green contours in **A-C** represent manual and automatic segmentation. Both versions of ResectVol (**A** and **B**) were applied to temporal and extratemporal data (n = 48), whereas DeepResection (**C**) is only applied to temporal cases (n = 23) due to its limitation. In **D** we display contours obtained with RVDL (red), RV112 (green), and DR (cyan) in temporal cases (to allow visual comparison across the three tools). Although included in boxplots and histograms, cases with DSC = 0 are not shown in the figures.

### 3.6. Large vs. Small lacunas

The Spearman’s correlation between volume (in cm³) and DSCs was r_s_ (46) = .60 (p < .001). To further assess this significant relationship, we ranked lacunas by their size obtained with the manual delineation and split our dataset into two subgroups based on the median lacuna volume (25.5 cm³). We investigated whether there was any difference in DSC across the small (< 25.5 cm³) and the large lacunas (> 25.5 cm³) (**Fig. 4**). Since the data in each group was not normally distributed, we performed a non-parametric Mann-Whitney U test that revealed a significant difference between groups (U = 450.5, p = .001).

**Fig. 4.**
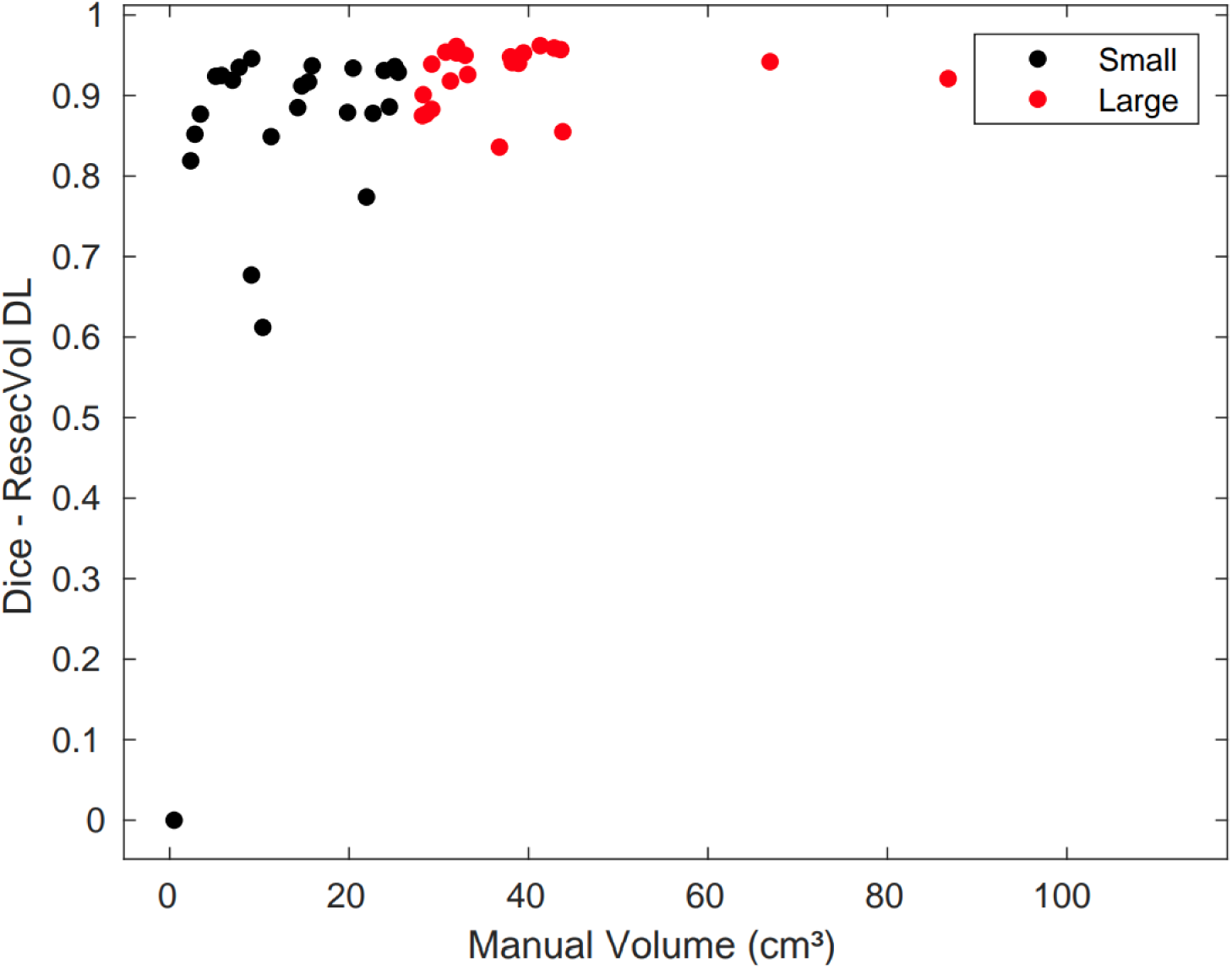
Distribution of Dice similarity coefficients (DSC) as a function of lacuna volume. Small lacunas (black dots) show more diverse values of DSC than large ones (red dots).

### 3.7. Structure Identification

After segmenting the lacuna, ResectVol DL combines the lacuna mask with the brain-extracted image to estimate the original 3D brain shape. This recreated whole brain undergoes a series of registration steps that register a labeled template in the Montreal Neurological Institute (MNI) space onto the space of the original image (native). We use the Desikan-Killiany-Tourville (DKT) atlas (19) as a reference for naming brain structures. Our pipeline uses functions from the Advanced Normalization Tools (ANTs) package (20), the Statistical Parametric Mapping (SPM) toolbox (21), and the FMRIB Software Library (FSL) (18). **Fig. 5** illustrates an example of the labeled regions of a temporal patient along with the table containing the volumetric information.

**Fig. 5.**
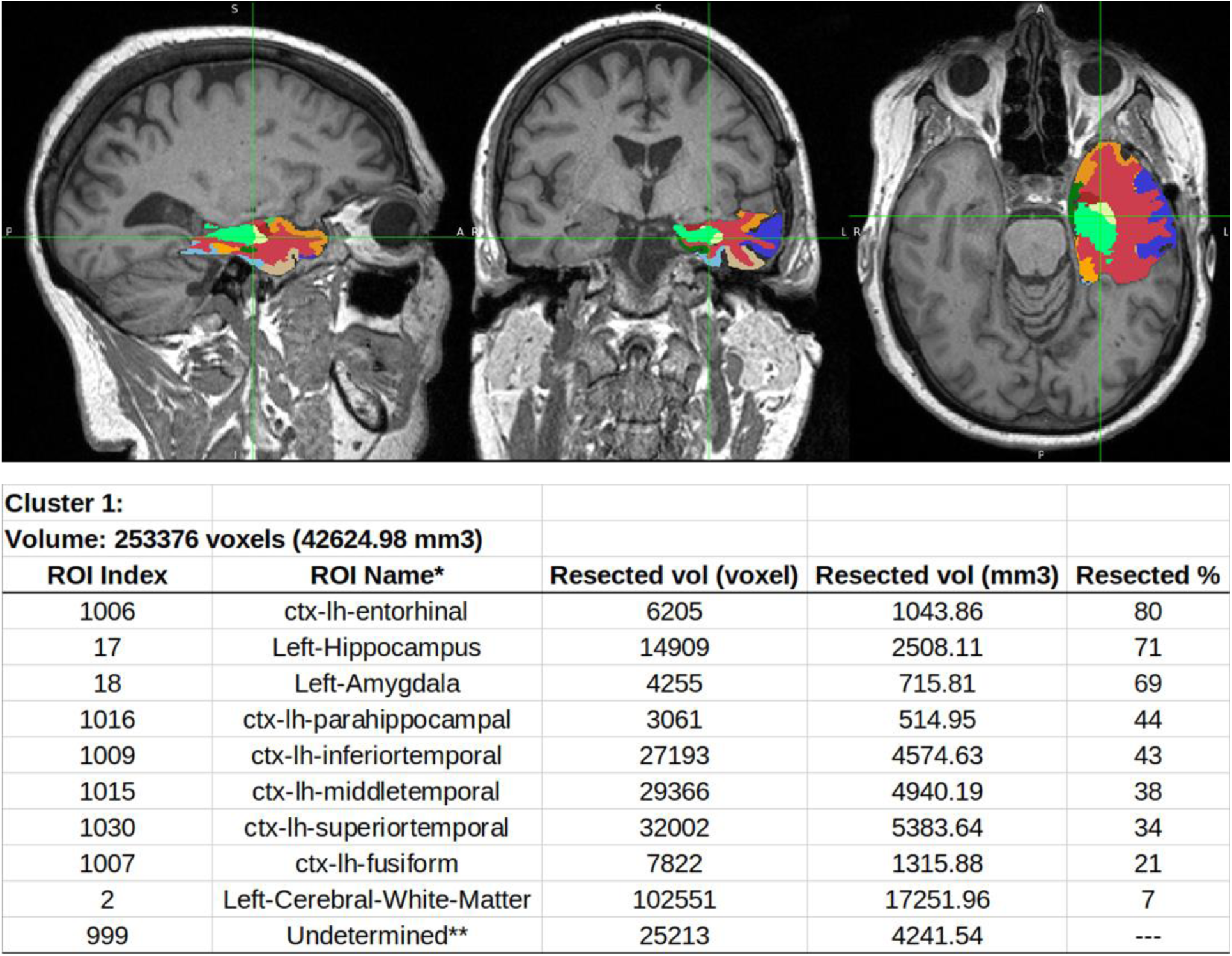
Region labeling in the lacuna segmentation (top) and the corresponding volumetric information table (bottom).

## 4. Discussion

In this study, we compared three different segmenting methods against the manual delineation of brain lacunas in T1-weighted MR images. We found that ResectVol DL, which relies on the nnU-Net architecture, could outperform its previous rule-based implementation (version 1.1.2) and DeepResection, a deep learning method based on the 2D U-Net architecture. To the best of our knowledge, DeepResection, ResectVol 1.1.2, and ResectVol DL are the only freely available, fully automatic tools dedicated to this task.

Lacuna segmentation can offer a means of estimating characteristics of the removed brain region and informing predictive models of surgery outcome. Manual segmentation is the obvious first approach considered, and arguably the one with the best quality, but it is susceptible to human error and consistency factors, such as inter and intra-rater variability. A subset of images from our study (n = 48) were segmented by two (or three) raters to estimate the inter-rater DSC. When more than two segmentations were available, we chose the highest associated DSC, yielding a more human-favorable estimate (DSC = .88) to be compared against the automatic methods.

Semi-automated methods have been implemented in which users need to click on a point inside the lacuna to carry out the segmentation. Gau et al. (22), for instance, employed itk-SNAP segmenting software (23) to perform the lacuna delineation by manually setting a seed inside the lacuna, followed by the itk-SNAP’s region-growing algorithm, and found a median DSC = .78 (range: .53 - .94). Billardello and colleagues (11) created a tool that also requires the user to click on (or set the coordinates of) a voxel to be used as the seed for the region-growing algorithm. They found a median DSC = .83 (Q1-Q3: .72 - .85). The major advantage of these approaches is that the seed is always positioned in the right location where the lacuna is expected to be identified. Nevertheless, this approach still demands human intervention, is prone to bias—since manual positioning of the seed may lead to different results—, and, although we did not make a direct comparison on the same dataset, the DSC found for ResectVol DL (.93) is better than in these studies.

Among the fully automatic tools we tested, DeepResection was the fastest (∼35 s per image), ResectVol DL was second (∼42 s), whereas ResectVol 1.1.2 took much longer (∼9 min) due to the nature of its processing algorithm which is based on a series of processing steps. When comparing median DSCs for temporal cases, ResectVol DL was the best one (DSC = .93) followed by DeepResection (.90) and ResectVol 1.1.2 (.79). Although DeepResection is capable of being fine-tuned to work with extratemporal resections—as tested in the original publication (exploratory DSC=.75)—authors do not recommend using this preliminary fine-tuned model. Hence, for extratemporal cases, we only considered the results from ResectVol 1.1.2 and ResectVol DL, and, again, version DL was the winner (.92 vs .73).

As previously reported in other studies (13,22), we found a relationship between DSC and lacuna size indicating that there is a trend towards better performance for larger resection. Since DSC is dependent upon a volume ratio, large values in the numerator (intersection) and denominator (total volume) will be less sensitive to intersection mismatches when comparing two volumetric shapes. Hence, small lacunas are more prone to be affected by discrepancies and present more variability in DSCs (**Fig. 4**).

Besides the volumetric information obtained with such computational segmentation methods, this line of research may offer potential avenues for surgery planning and outcome prediction. Koepp et al. (24) showed that the removal of at least 50 % of the piriform cortex increased (by a factor of 16) the odds of becoming seizure-free, and no relationship was found with other mesiotemporal structures. Leon-Rojas and colleagues (25) implemented a pipeline, based on this result and on their dataset of resective surgeries, to automatically segment the piriform cortex and calculate chances of seizure freedom during planning. They were able to plan a new patient intervention with an estimated 50 % chance of long-term seizure freedom. The automatic segmentation of resected areas can help increase knowledge and accelerate the development of such tools. Since ResectVol can list areas removed along with their volume, it can also help serve that purpose.

Regarding manual segmentation, raters spent more time in our study (approximately or more than 1 hour) than what has been reported by other authors (approximately 30 minutes) (12,22). We found that duration depends on lesion complexity (as reported in these studies), tool used, and rater experience.

Computer systems can spare humans from these repetitive tasks and accelerate their completion, besides avoiding the mentioned human error and variability. In the inter-rater assessment, we found a DSC = .88, comparable to previous reports on brain tumors (.84 - .86) (26) and lacuna resection (.84) (27). We noticed idiosyncrasies among raters, that were more pronounced in temporal resections, especially in the very medial region near the hippocampus. We do recommend that researchers carefully check, or standardize—if possible—, important resection areas if they anticipate that some regions may be controversial.

One limitation of our tool refers to not being tested on postoperative MRIs of other surgery types, like laser interstitial thermal therapy. However, we anticipate that, due to the different profiles of lesioned brain sites, a new network should be trained on this specific dataset to accommodate these discordant characteristics. We also have not validated the labeling of resected brain structures, which is an area that does deserve attention in future studies. Overall, ResectVol DL had the best performance across the tested tools, having failed for only one case out of 53 on which it was tested. It can be utilized in temporal and extratemporal cases and is freely available at github.com/rfcasseb/resectvol_dl/.

## 5. Conclusion

ResectVol DL successfully segmented brain lacunas in MRIs of patients with epilepsy who have undergone surgery. ResectVol DL is fast, accurate, and does not require any image pre-processing. It relies solely on postoperative images to measure lacuna volumes besides providing estimates of the volumes of anatomical structures within the lacuna. This enables more precise estimation of volumetric information, which facilitates studies such as fMRI investigations performed before surgery. Moreover, it may aid in generating predictive models of surgical results.

### Ethics statement

This study was conducted with approval from the Cleveland Clinical Foundation Institutional Review Board. Informed consent was waived due to the retrospective nature of the data collection.

## Data Availability

The data that support the findings of this study can be made available after approval from the ethics committee and through a formal data sharing agreement.

## Acknowledgements

This work was supported by the São Paulo Research Foundation [grant numbers 2020/00019-7 and 2013/07559-3] and the National Institutes of Health [grant number R01NS097719].

## CRediT authorship contribution statement

**Raphael Fernandes Casseb:** Conceptualization, Methodology, Software, Data Curation, Formal Analysis, Visualization, Writing – original draft, Writing – review & editing. **Brunno Machado de Campos:** Conceptualization, Methodology, Software, Formal Analysis, Writing – original draft, Writing – review & editing, **Wallace Souza Loos:** Methodology, Software, Writing – review & editing. **Marcelo Eduardo Ramos Barbosa:** Data Curation, Formal Analysis, Writing – review & editing. **Marina Koutsodontis Machado Alvim**: Data Curation, Investigation, Writing – review & editing, **Gabriel Chagas Lutfala Paulino:** Data Curation, Formal Analysis, Writing – review & editing. **Francesco Pucci**: Data Curation, Formal Analysis, Writing – review & editing. **Samuel Worrell**: Data Curation, Formal Analysis, Writing – review & editing. **Roberto Medeiros de Souza:** Methodology, Software, Writing – review & editing. **Lara Jehi:** Conceptualization, Resources, Project administration, Supervision, Writing – review & editing. **Fernando Cendes:** Conceptualization, Supervision, Resources, Project administration, Writing – original draft, Writing – review & editing.

